# Diagnostic utility of plasma ptau217, ptau181, GFAP for Alzheimer disease in a heterogeneous younger onset dementia clinical cohort

**DOI:** 10.1101/2024.04.29.24306586

**Authors:** Dhamidhu Eratne, Qiao-Xin Li, Courtney Lewis, Christa Dang, Matthew JY Kang, Jasleen Grewal, Samantha Loi, Mark Walterfang, Andrew H Evans, Charles B Malpas, Steve Pedrini, Ralph Martins, Pratishtha Chatterjee, Henrik Zetterberg, Kaj Blennow, Samuel F Berkovic, Alexander F Santillo, Steven Collins, Colin L Masters, Dennis Velakoulis, The MiND Study Group

**Affiliations:** Neuropsychiatry, Royal Melbourne Hospital; Melbourne Neuropsychiatry Centre & Department of Psychiatry, University of Melbourne; National Dementia Diagnostics Laboratory, The Florey, Melbourne; National Dementia Diagnostics Laboratory (NDDL), The Florey Institute, The University of Melbourne, 30 Royal Parade, Parkville VIC 3010, Australia; Department of Psychiatry, University of Melbourne; Institute of Health and Wellbeing Federation University; National Ageing Research Institute; Department of General Practice, University of Melbourne; Neuropsychiatry Centre, Royal Melbourne Hospital; Department of Psychiatry, University of Melbourne; The Alfred Hospital, Melbourne, Australia; Neuropsychiatry Centre, RMH and Department of Psychiatry, University of Melbourne; Neuropsychiatry Centre, Royal Melbourne Hospital, Melbourne, Australia; Department of Psychiatry, University of Melbourne, Melbourne, Australia; Florey institute of Neuroscience and Mental Health, Melbourne, Australia; Edith Cowan University, Perth, Australia; Neuropsychiatry Centre, Royal Melbourne Hospital, Department of Medicine, University of Melbourne, Epworth Hospital; Department of Medicine, Royal Melbourne Hospital, University of Melbourne, Melbourne, Australia; Melbourne School of Psychological Sciences, University of Melbourne, Melbourne, Australia; Edith Cowan University, Joondalup, 6027; Alzheimers Research Australia, Nedlands 6009; Edith Cowan University, Perth Australia & Macquarie University, Macquarie Medical School, Sydney Austraila; The Florey Institute, The University of Melbourne, 30 Royal Parade, Parkville VIC 3052, Australia; ^1^Department of Psychiatry and Neurochemistry, Institute of Neuroscience and Physiology, the Sahlgrenska Academy at the University of Gothenburg, Mölndal, Sweden; ^2^Clinical Neurochemistry Laboratory, Sahlgrenska University Hospital, Mölndal, Sweden; ^3^Department of Neurodegenerative Disease, UCL Institute of Neurology, Queen Square, London, UK; ^4^UK Dementia Research Institute at UCL, London, UK; ^5^Hong Kong Center for Neurodegenerative Diseases, Clear Water Bay, Hong Kong, China; ^6^Wisconsin Alzheimer’s Disease Research Center, University of Wisconsin School of Medicine and Public Health, University of Wisconsin-Madison, Madison, WI, USA; Institute of Neuroscience and Physiology, University of Gothenburg, Mölndal, Sweden; Clinical Neurochemistry Lab, Sahlgrenska University Hospital, Mölndal, Sweden; Paris Brain Institute, ICM, Pitié Salpêtrière Hospital, Sorbonne University, Paris, France; Neurodegenerative Disorder Research Center, Division of Life Sciences and Medicine, and Department of Neurology, Institute on Aging and Brain Disorders, University of Science and Technology of China and First Affiliated Hospital of USTC, Hefei, P.R. China; Epilepsy Research Centre, Department of Medicine, Austin Health, The University of Melbourne, Heidelberg; Department of Clinical Sciences, Clinical Memory Research Unit, Faculty of Medicine, Lund University, Malmo, Sweden; Department of Medicine (RMH) and National Dementia Diagnostics Laboratory, The Florey, The University of Melbourne; National Dementia Diagnostics Laboratory, The Florey, The University of Melbourne; Neuropsychiatry, Royal Melbourne Hospital; Melbourne Neuropsychiatry Centre & Department of Psychiatry, University of Melbourne

## Abstract

**Objective:** We investigated diagnostic utility of phosphorylated tau 217 and 181 (ptau217, ptau181), glial fibrillary acidic protein (GFAP), amyloid beta 42 and 40 (Aβ42, Aβ40), neurofilament light (NfL) to distinguish AD from non-AD conditions, in a heterogenous clinical cohort of younger people.

**Methods:** Plasma biomarkers were analysed using ultrasensitive technology, and compared in patients with CSF Alzheimer disease profiles (A+T+) to other profiles (OtherAT).

**Results:** Seventy-nine patients were included, median age 60.8 years: 16 A+T+, 63 OtherAT. Ptau217, ptau181, GFAP were significantly elevated in A+T+ compared to OtherAT (3.67 vs 1.12pg/mL, 3.87 vs 1.79pg/mL, 189 vs 80pg/mL, respectively). ptau217 distinguished AD from OtherAT with 90% accuracy (88% specificity, 100% sensitivity)

**Conclusions:** Plasma ptau217 has strong diagnostic utility to diagnose AD in a clinically relevant, younger cohort of people with symptoms, adding further weight for a simple diagnostic blood test for AD as a cause of a patient’s symptoms.

## INTRODUCTION

In recent years there have been significant developments in identifying blood biomarkers which can provide simple, accurate, widely available, and easily accessible diagnostic tests for people with dementia. Such blood-based diagnostic biomarkers could markedly increase worldwide access and adoption, with significant positive impacts for individual patients, their families, but also with wider healthcare systems and public health implications [1,2]. Two promising diagnostic biomarkers for Alzheimer disease have been phosphorylated tau 217 (ptau217) and phosphorylated tau 181 (ptau181), with the former holding promise to be the ‘simple blood test for Alzheimer disease’ [3–9]. Other promising biomarkers include glial fibrillary acidic protein (GFAP) in AD and other disorders, [6,10] and neurofilament light chain (NfL) in neurodegenerative disorders broadly and in distinguishing neurodegenerative from non-neurodegenerative and neuroinflammatory disorders. [11,12,12,13].

The focus of most research has been on AD and the AD continuum, and in older people. There has been limited literature on plasma ptau217 and other AD biomarkers in clinically heterogeneous cohorts (including AD as well as non-AD neurodegenerative disorders and primary psychiatric conditions), and limited research in younger people with dementia (traditionally defined in dementia care and research as symptom onset under 65 years of age). However, real-world clinical practice and research in younger onset dementia services and settings has consistently demonstrated the significant diagnostic challenges, broader range of diagnoses encountered (including non-AD neurodegenerative disorders such as frontotemporal dementia, and primary psychiatric disorders), uncertainty, misdiagnosis and diagnostic delay in younger people, despite access to current gold standard assessments [13–18]. Furthermore, most studies in research settings have excluded participants with significant psychiatric and other comorbidities, limiting generalisability. It is imperative to understand the performance of blood-based biomarkers in such clinically relevant and generalisable, heterogenous cohorts across the lifespan to properly understand the strengths and limitations, and help ensure the correct implementation of a widely available blood test for AD.

The primary aim of this study was to investigate blood biomarker levels (ptau217, ptau181, GFAP, amyloid beta Aβ42, Aβ40, NfL) and their diagnostic utility for AD versus non-AD, in a heterogenous clinical cohort of younger people with symptoms and presenting for diagnostic assessment at a specialist young onset dementia service.

For the purpose of this study, we defined AD based on classification of cerebrospinal fluid (CSF) AD proteins, recommended for diagnosis [19], and which have been validated against amyloid positron emission tomography (PET; currently considered ‘gold standard’ for a premortem diagnosis of AD) with over 90% concordance [20–22]. The CSF AD biomarkers, amyloid beta 1-42 (Aβ42), a component of amyloid plaques, and phosphorylated tau (ptau), the main component of neurofibrillary tangles, are clinically validated diagnostic tests for AD, and are routinely used in many centres to discriminate AD from other causes of dementia in people with symptoms presenting to clinical services for diagnostic assessment. Based on and consistent with the AT(N) framework definition of AD [23], we therefore defined AD based on a positive amyloid and positive phosphorylated (A+T+) CSF profile, similar to our recent study and other studies.

Other CSF profiles are not considered diagnostic of AD nor the cause of patients’ symptoms. An isolated low CSF amyloid and normal p-tau level (A+T-profile) is a common non-specific finding in other, non-AD neurodegenerative conditions such as frontotemporal dementia, vascular dementia, dementia with Lewy bodies, and even in primary psychiatric disorders.[24,25]. Furthermore, even if a profile such as A+T-might be consistent with or suggest “Alzheimer’s pathological change” or preclinical Alzheimer’s continuum, this is not considered the cause when a patient is symptomatic and presenting to specialist services. We therefore defined CSF profiles other than A+T+ as “OtherAT”.

The primary hypothesis was that levels of AD blood biomarkers ptau217, ptau181, and GFAP would be higher in A+T+ compared to OtherAT (A-T-, A-T+, A+T-) CSF profiles, and that these biomarkers would have high diagnostic utility in distinguishing between A+T+ from OtherAT in a clinical cohort of people with symptoms in a specialist service. We hypothesised that NfL would not distinguish between A+T+ and OtherAT given its lack of specificity for specified neurodegenerative disorders, and since the OtherAT group would include both neurodegenerative disorders associated and non-neurodegenerative disorders. Secondary aims were to investigate the levels and diagnostic utility between different CSF profiles.

## METHODS

### Study cohort

Patients were recruited from the Neuropsychiatry Centre, The Royal Melbourne Hospital, Australia. As previously described [12,13,17], The Neuropsychiatry Centre is a tertiary clinical service providing diagnostic input to people with a range of neuropsychiatric presentations, particularly young-onset dementia. Patients are referred for diagnostic assessment and management of possible young-onset dementia and undergo comprehensive multidisciplinary and multimodal assessments and investigations, including CSF AD proteins. Patients were included in this study if they had blood biomarker analyses and CSF AD protein analysis within 12 months.

### Biomarker analyses

Plasma ptau217 was analysed using the University of Gothenburg (UGOT) ptau217 assay, as previously described [7]. Other plasma biomarkers were analysed using the Quanterix Simoa HD-X platform. NfL and GFAP were analysed using the 2-plex assay. AB42 and AB40 data was available from analysis using the N4PE assay, and ptau181 using the single plex assay.

CSF biomarkers were analysed for clinical diagnostic purposes using Roche Elecsys and INNOTEST ELISA platforms. Amyloid beta positivity was determined based on established cut-offs (<1030pg/mL for Roche Elecsys platform, < 656pg/mL for INNOTEST platform). The established cutoffs for ptau181 positivity were >27pg/mL (Elecsys), and for INNOTEST >=59 (<=70 year olds) and >=74 (>70 year olds). CSF profiles were categorised based on the beta amyloid and ptau status, based on the AT(N) framework: A-T-, A-T+, A+T-, A+T+. An A+T+ CSF profile was considered consistent with AD as defined and described earlier, and other AT profiles were groups as ‘OtherAT’, in keeping with biomarker definitions of AD [23,26–28] and described further in our recent study [29].

### Statistical analyses

Statistical analyses were performed using R version 4.3.2 (2023-10-31). As several biomarker distributions were non-Gaussian (even when log transformed), biomarker levels in different groups were compared using standardised bootstrapped quantile regression, with age and sex as additional covariates. ROC curve analyses were performed to investigate diagnostic utility between different combinations of groups.

This study, part of The Markers in Neuropsychiatric Disorders Study (The MiND Study, https://themindstudy.org), was approved by the Human Research Ethics Committees at Melbourne Health (2016.038, 2017.090, 2018.371, 2020.142), University of Melbourne (1341074), St. Vincent’s Hospital (028-06), and the Florey Institute of Neurosciences and Mental Health (1648441.1).

## RESULTS

The final cohort included 79 patients, 16 with AD (A+T+ CSF profile), 63 with OtherAT (A-T-, A-T+, A+T-CSF profiles). As demonstrated in Table 1, this was a younger cohort with no difference in age between A+T+ and OtherAT groups (median age 61.4 vs 60.2 years). There was a higher proportion of females in the A+T+ group (63% vs 35%). All 79 patients had recorded GFAP and NfL levels, and 73 had recorded ptau217 levels. 63 patients had recorded ptau181, AB42, and AB40 levels.

**Table 1.** Demographics and plasma biomarkers in CSF Alzheimer disease profile (A+T+) versus Other CSF profile groups. Data are median [interquartile range] or n (%). All biomarker levels are in pg/mL. AB40 = amyloid beta 40; AB42 = amyloid beta 42; GFAP = glial fibrillary acidic protein; ptau181 = phosphorylated tau 181; ptau217 = phosphorylated tau 217; NfL = neurofilament light chain protein

|  | [ALL]<br>N=79 | OtherAT<br>N=63 | A+T+<br>N=16 | N |
| --- | --- | --- | --- | --- |
| Age | 60.8 [57.1;64.7] | 60.2 [57.1;64.2] | 61.4 [58.5;64.9] | 79 |
| Sex: Female | 32 (40.5%) | 22 (34.9%) | 10 (62.5%) | 79 |
| ptau217 | 1.37 [0.82;2.61] | 1.12 [0.73;1.58] | 3.67 [3.17;4.60] | 73 |
| ptau181 | 2.12 [1.26;3.14] | 1.79 [1.18;2.65] | 3.87 [2.45;4.22] | 63 |
| GFAP | 128 [73.5;216] | 114 [66.1;174] | 224 [185;262] | 79 |
| Aβ42 | 5.80 [4.96;6.75] | 5.87 [4.87;6.77] | 5.59 [5.08;6.14] | 63 |
| Aβ40 | 95.8 [85.4;115] | 93.4 [85.2;113] | 104 [92.5;117] | 63 |
| Aβ42: Aβ40 | 0.06 [0.05;0.06] | 0.06 [0.05;0.07] | 0.06 [0.05;0.06] | 63 |
| NfL | 17.9 [11.1;28.5] | 16.9 [10.3;31.8] | 21.2 [15.8;26.8] | 79 |

Reflective of the clinical setting and cohort, patients in the OtherAT group had a diverse range of clinical diagnoses, including non-Alzheimer neurological and neurodegenerative disorders, mild cognitive impairment, and primary psychiatric disorders.

### Biomarker levels and diagnostic utility in A+T+ vs OtherAT

As demonstrated in Table 1 and Figure 1, plasma ptau217 levels were significantly elevated in A+T+ compared to OtherAT (median 3.67pg/mL vs 1.12pg/mL); standardised quantile regression coefficient, ß: 1.64, 95%CI [1.27, 2.14], p < 0.001.

**Figure 1.**
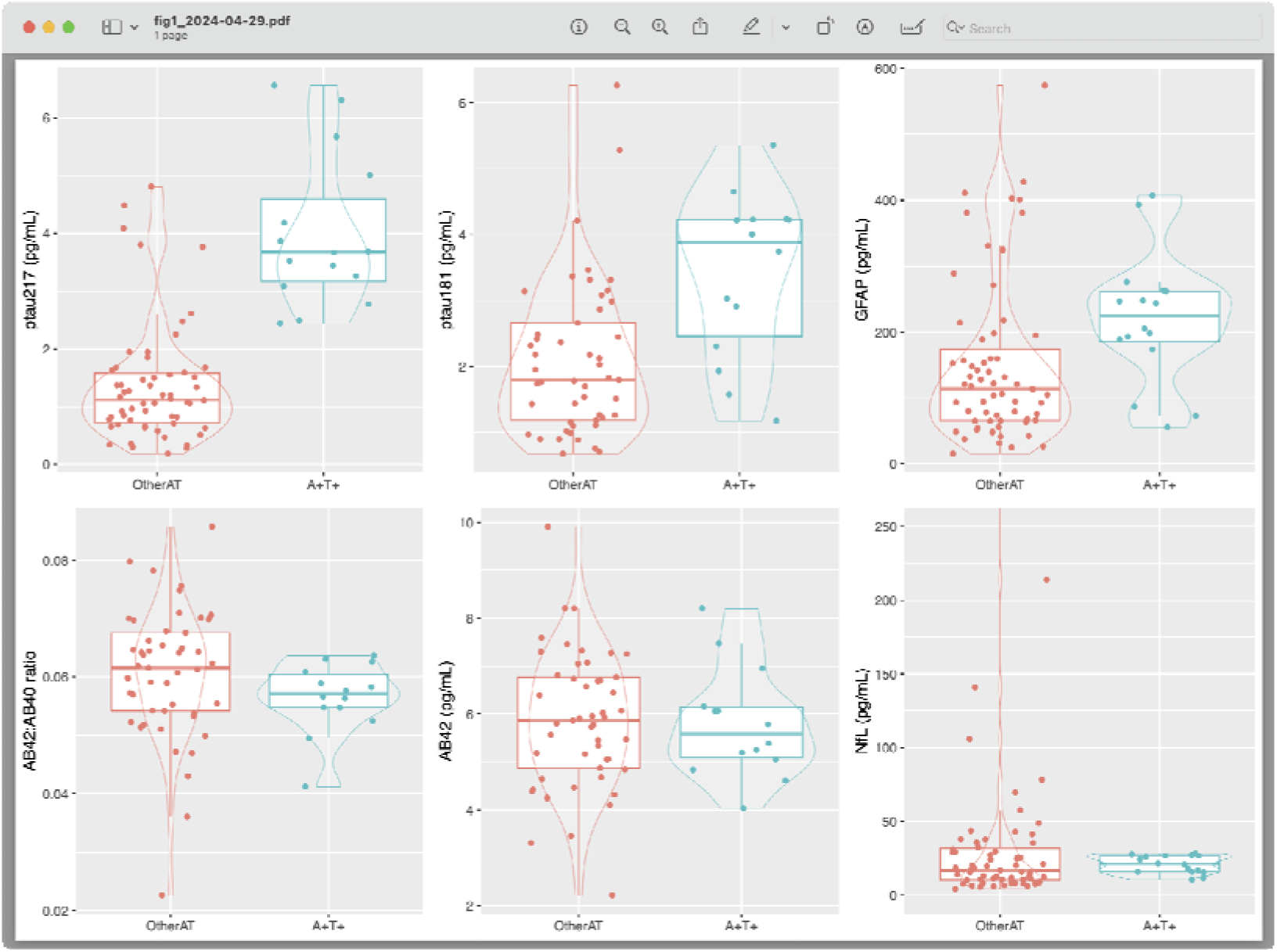
Plasma biomarker levels in CSF A+T+ profiles compared to Other profiles. To improve readability of NfL plot, an extreme outlier (1655pg/mL) is not shown on the plot. AB40 = amyloid beta 40; AB42 = amyloid beta 42; GFAP = glial fibrillary acidic protein; ptau181 = phosphorylated tau 181; ptau217 = phosphorylated tau 217; NfL = neurofilament light chain protein

Plasma ptau181 and GFAP levels were also elevated in A+T+ compared to OtherAT (3.87 vs 1.79 and 189 vs 80, respectively), with smaller effect sizes (ß: 1.07, 95%CI [0.18, 1.82], p=0.014, and ß: 0.85, 95%CI [0.39, 1.35], p=0.014, respectively). In contrast, there was no statistical evidence for differences in Aβ42:Aβ40 ratio and NfL levels between AD and OtherAT (ß: -0.57, 95%CI [-1.14, 0.10], p=0.098, and ß: -0.02, 95%CI [-0.04, 0.32], p=0.302, respectively).

In terms of diagnostic utility (Figure 2), Plasma ptau217 had the strongest performance to distinguish AD from OtherAT, with an area under the curve (AUC) of 0.94 [0.89, 0.99], and an optimal cut-off of 2.35pg/mL, associated with 88% specificity, 100% sensitivity, positive likelihood ratio (LR+) 8.29, negative likelihood ratio (LR-) of 0, 68% positive predictive value (PPV), 100% negative predictive value (NPV), and 90% accuracy.

**Figure 2.**
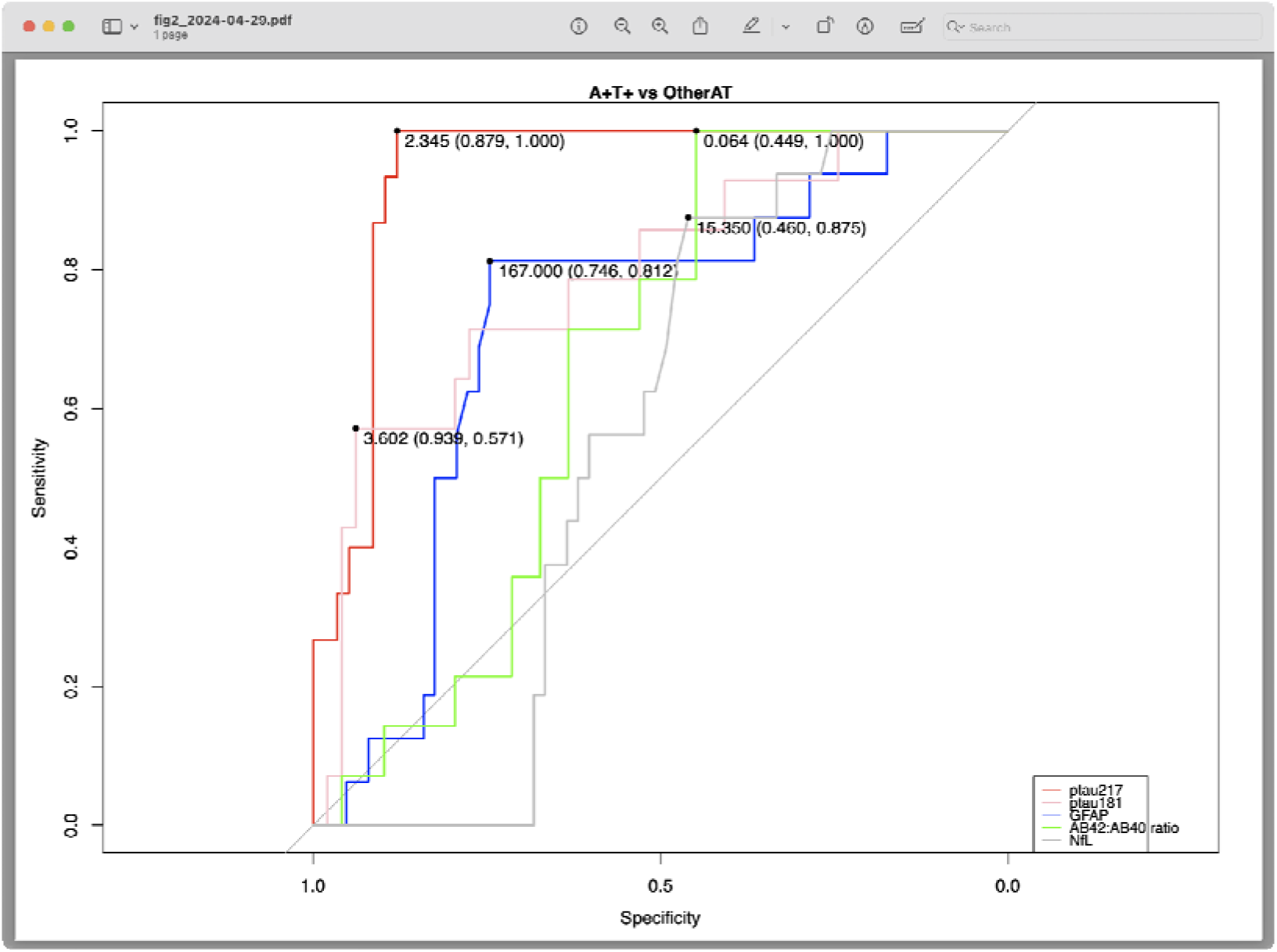
Receiver operator curve analysis for plasma biomarkers to disinguish CSF A+T+ from OtherAT CSF profiles. Numbers on figure are optimal cut-off (specificity, sensitivity). AB40 = amyloid beta 40; AB42 = amyloid beta 42; GFAP = glial fibrillary acidic protein; ptau181 = phosphorylated tau 181; ptau217 = phosphorylated tau 217; NfL = neurofilament light chain protein

Results for the other biomarkers were, ptau181: AUC 0.79 [0.65, 0.93], cut-off 3.6pg/mL, 94% specificity, 57% sensitivity, LR+ 9.33, LR-0.46, PPV 73%, NPV 88%, 86% accuracy; GFAP AUC 0.72 [0.58, 0.86], cut-off 167, 75% specificity, 81% sensitivity, LR+ 3.2, LR-0.25, PPV 45%, NPV 94%, 76% accuracy; Aβ42:Aβ40 AUC 0.65 [0.52, 0.79], cut-off 0.064, 44% specificity, 100% sensitivity, LR+ 1.81, LR-0, PPV 34%, NPV 100%, accuracy 57%. NfL did not have diagnostic utility (AUC 0.56 [0.44, 0.68]).

### Biomarker levels and diagnostic utility between specific AT groups

Plasma ptau217 levels were significantly higher in A+T+ compared to A-T-(3.67pg/mL vs 1.05pg/mL; ß: 1.71 [1.35, 2.24], p<0.001) and A+T-(3.67 vs 1.30pg/mL, ß: 1.54 [1.12, 2.11], p<0.001), but not A-T+ (p=0.154), Table 2 and Figure 3.

**Table 2.** Demographics and plasma biomarkers in different CSF profiles. Data are median [interquartile range] or n (%). AB40 = amyloid beta 40; AB42 = amyloid beta 42; GFAP = glial fibrillary acidic protein; ptau181 = phosphorylated tau 181; ptau217 = phosphorylated tau 217; NfL = neurofilament light chain protein

|  | A-T-<br>N=26 | A-T+<br>N=6 | A+T-<br>N=31 | A+T+<br>N=16 | N |
| --- | --- | --- | --- | --- | --- |
| Age | 58.4 [56.3;62.4] | 62.5 [60.2;63.8] | 61.0 [57.1;65.3] | 61.4 [58.5;64.9] | 79 |
| Sex: Female | 12 (46.2%) | 2 (33.3%) | 8 (25.8%) | 10 (62.5%) | 79 |
| ptau217 | 1.05 [0.70;1.23] | 1.63 [0.96;1.68] | 1.30 [0.77;1.92] | 3.67 [3.17;4.60] | 73 |
| ptau181 | 1.73 [1.16;1.98] | 3.14 [2.99;3.31] | 2.17 [1.42;2.86] | 3.87 [2.45;4.22] | 63 |
| GFAP | 80.0 [65.2;133] | 85.8 [52.7;141] | 140 [94.1;254] | 224 [185;262] | 79 |
| Aβ42 | 5.95 [5.26;7.17] | 7.28 [4.68;7.32] | 5.78 [4.87;6.44] | 5.59 [5.08;6.14] | 63 |
| Aβ40 | 91.9 [82.3;111] | 118 [93.8;128] | 91.7 [85.2;109] | 104 [92.5;117] | 63 |
| Aβ42: Aβ40 | 0.06 [0.06;0.07] | 0.05 [0.05;0.06] | 0.06 [0.05;0.07] | 0.06 [0.05;0.06] | 63 |

**Table 2. Demographics and plasma biomarkers in different CSF profiles.**
|  | A-T- | A-T+ | A+T- | A+T+ | N |
| --- | --- | --- | --- | --- | --- |
|  | N=26 | N=6 | N=31 | N=16 |  |
| NfL | 19.4 [8.46;37.3] | 16.9 [11.5;35.6] | 15.8 [11.1;26.4] | 21.2 [15.8;26.8] | 79 |

**Figure 3.**
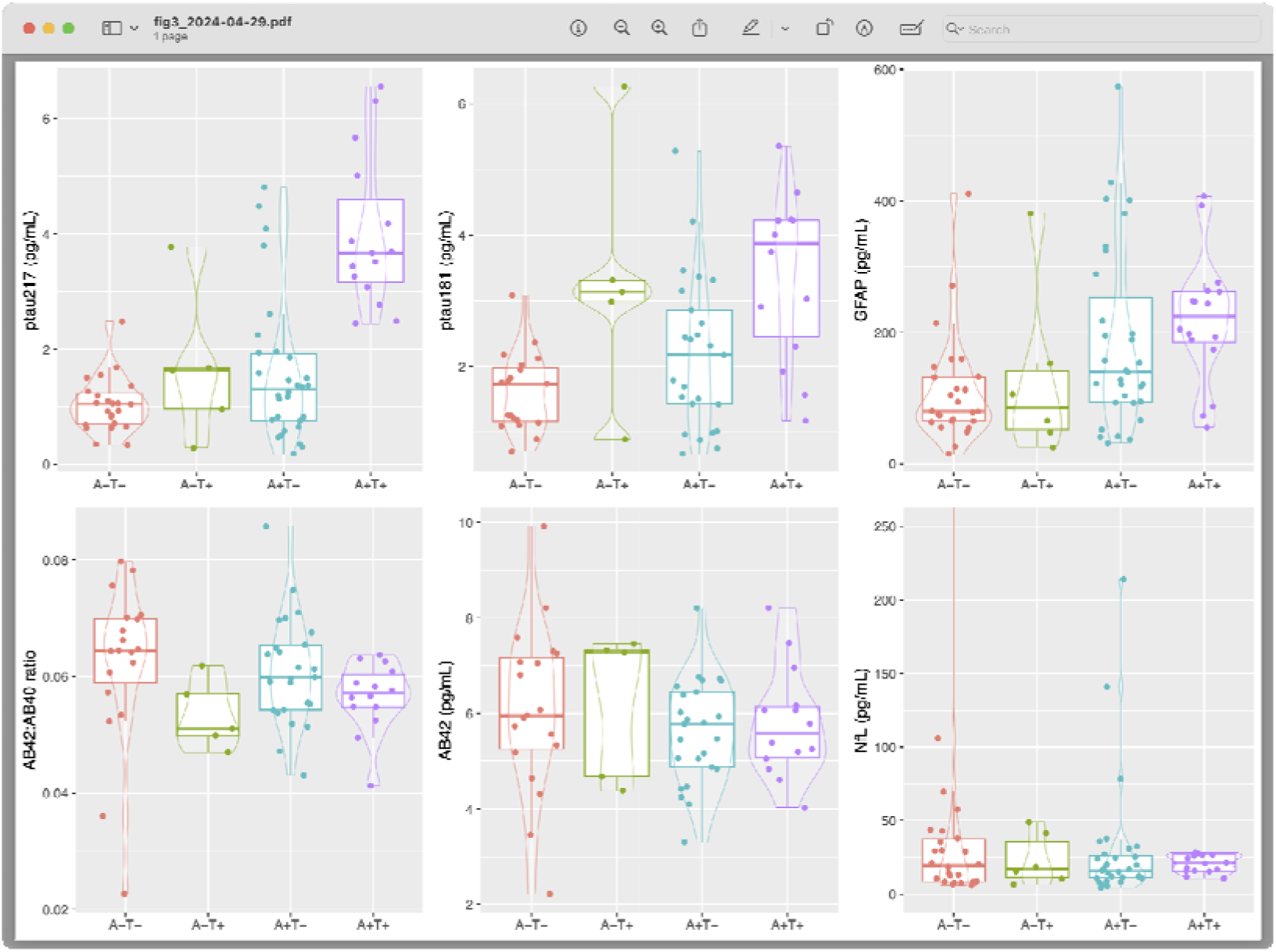
Plasma biomarker levels in different CSF profiles. To improve readability of NfL plot, an extreme outlier (1655pg/mL) is not shown on the plot. AB40 = amyloid beta 40; AB42 = amyloid beta 42; GFAP = glial fibrillary acidic protein; ptau181 = phosphorylated tau 181; ptau217 = phosphorylated tau 217; NfL = neurofilament light chain prote

Ptau181 levels were higher in A+T+ compared to A-T-(3.87pg/mL vs1.73pg/mL; ß: 1.32 [0.32, 2.01], p=0.012), and A+T-(3.87 vs 2.17; ß: 1.02 [0.04, 1.91], p=0.038), but not A-T+ (p=0.748). GFAP was elevated in A+T+ compared to A-T-(224 vs 80; ß: 0.95 [0.44, 1.49], p=0.006), but not A-T+ (p=0.134), or A+T-(p=0.094). Aβ42:Aβ40 ratio was slightly lower in A+T+ compared to A-T-(ß: -0.87 [-1.36, - 0.20], p=0.008, but not different to the other groups (A-T+ p=0.781, A+T-p=0.137). NfL levels were not different between any of the groups (all p>0.27).

Ptau217 distinguished A+T+ versus A-T-with high accuracy (AUC 0.997 [0.99, 1], cut-off 2.06pg/mL, 96% specificity, 100% sensitivity), as well as A+T+ vs A+T-(AUC 0.90 [0.81, 1], cut-off 2.35, 83% specificity, 100% sensitivity), and A+T+ vs A-T+ (AUC 0.88 [0.64, 1], cut-off 2.06, 80% specificity, 100% sensitivity). Ptau217 was superior to ptau181 and other biomarkers. Full details are available in Supplementary Material.

### Sensitivity analyses

Several sensitivity analyses were performed. One patient had an extremely high NfL level (1655pg/mL). Analyses excluding this extreme outlier and restricting the sample to those with blood and CSF samples being closer together (being within 6 months of each other, and being within 3 months of each other), produced very similar results.

## DISCUSSION

This study found significantly elevated plasma ptau217 levels in patients with AD as defined by an A+T+ CSF profile, thereby establishing strong diagnostic utility of ptau217 for AD, in younger patients with a broad range of cognitive, psychiatric, neurological symptoms. This builds the evidence of the utility of a blood test to diagnose AD, especially in a younger, diagnostically heterogenous, and clinically relevant cohort, which is of particular importance given the relative lack of research in this group that has higher rates of diagnostic delay and misdiagnosis. In addition, this study provides evidence of the superiority of ptau217 compared to ptau181 and other biomarkers to diagnose AD, and the poor ability of Quanterix Simoa Aβ42 and Aβ40 to assist in this distinction [30].

The study cohort was purely a clinical cohort of patients who were referred to and assessed for neuropsychiatric/cognitive symptoms at a specialist young onset dementia service, and were diagnosed with a broad range of neurodegenerative and primary psychiatric disorders based on comprehensive clinical assessments and investigations. While other studies did not focus on purely clinical cohorts, included only participants in the Alzheimer’s continuum, or only compared AD against healthy controls, the clinically relevant cohort and AT characterisation are strengths of this study. By examining a younger, heterogenous clinical cohort, this study provides data on real-world utility at the clinical coalface, given that the challenge in current real-world clinical practice is timely and accurate diagnosis of AD for people with cognitive and neuropsychiatric symptoms, not from distinguishing AD from healthy controls or from preclinical Alzheimer’s.

An important point is that the A-T-patients in this study include people with various neurodegenerative and primary psychiatric diagnoses, so this group is not comparable to AD-focused studies where the A-T-group is equivalent or almost equivalent to healthy controls. This would be the main explanation of our (expected) finding a lack of utility of NfL to distinguish from A+T+ from diverse non-AD conditions, given even the A-T-group included many people with a neurodegenerative disorder known to be associated with elevated NfL levels [12,31]. The findings of this study fit with existing work demonstrating promise in using NfL as a higher level or first tier test to distinguish neurodegenerative from non-neurodegenerative and primary psychiatric conditions [12,31], with more specific tests such as ptau217 being more appropriately used to distinguish AD from other neurodegenerative conditions. Identifying optimal biomarkers for such tiered testing is particularly useful as precision medicine approaches and diagnostic algorithms for timely and accurate diagnosis of dementia emerge.

Limitations include a lack of data on factors that have been shown to influence biomarker levels to a degree (weight, renal function, and whether samples were fasted or not). Despite this, the younger cohort and the high diagnostic utility of ptau217 means that the overall findings are less likely to be dramatically influenced by such variables. This study focused on a clinical cohort and did not have a control group for comparison. However, we could expect that healthy controls would be likely to have a narrower range or lower levels of ptau217, and therefore expect the diagnostic performance to be even stronger in distinguishing A-T-healthy controls from A+T+. While this cohort did not have amyloid PET, genetic, or pathological confirmation, CSF profiles have been shown to correlate strongly with amyloid PET, and CSF AD biomarker analysis is more widely accessible than amyloid PET. Finally, the relatively small numbers within subgroups based on AT status (especially the A-T+ group), mean that findings and interpretations need further study and validation in larger cohorts.

To conclude, this study demonstrated strong diagnostic utility of plasma ptau217 to distinguish AD from other CSF profiles in a clinically relevant, younger cohort of people with symptoms, adding further weight for a simple diagnostic blood test for AD as a cause of a patient’s symptoms. There is great promise of blood biomarkers to improve timely, accurate diagnosis, of particular importance in this younger group, where diagnostic delay, misdiagnosis, atypical presentations, wider differential diagnoses, are all more common [16,18]. A simple blood test for AD could have significant and far-reaching positive impacts for patients, families, and healthcare systems, especially in this era of disease-modifying treatments for AD.

## Data Availability

All data produced in the present study are available upon reasonable request to the authors

## ACKNOWLEDGEMENTS AND FUNDING SOURCES

HZ is a Wallenberg Scholar and a Distinguished Professor at the Swedish Research Council supported by grants from the Swedish Research Council (#2023-00356; #2022-01018 and #2019-02397), the European Union’s Horizon Europe research and innovation programme under grant agreement No 101053962, Swedish State Support for Clinical Research (#ALFGBG-71320), the Alzheimer Drug Discovery Foundation (ADDF), USA (#201809-2016862), the AD Strategic Fund and the Alzheimer’s Association (#ADSF-21-831376-C, #ADSF-21-831381-C, #ADSF-21-831377-C, and #ADSF-24-1284328-C), the Bluefield Project, Cure Alzheimer’s Fund, the Olav Thon Foundation, the Erling-Persson Family Foundation, Stiftelsen för Gamla Tjänarinnor, Hjärnfonden, Sweden (#FO2022-0270), the European Union’s Horizon 2020 research and innovation programme under the Marie Skłodowska-Curie grant agreement No 860197 (MIRIADE), the European Union Joint Programme – Neurodegenerative Disease Research (JPND2021-00694), the National Institute for Health and Care Research University College London Hospitals Biomedical Research Centre, and the UK Dementia Research Institute at UCL (UKDRI-1003). MK is supported by the Research Training Program Scholarship from the Department of Psychiatry, University of Melbourne with contributions from the Australian Commonwealth Government, and the Ramsay Hospital Research Foundation.

## Acknowledgements / Funding

KB is supported by the Swedish Research Council (#2017-00915 and #2022-00732), the Swedish Alzheimer Foundation (#AF-930351, #AF-939721, #AF-968270, and #AF-994551), Hjärnfonden, Sweden (#FO2017-0243 and #ALZ2022-0006), the Swedish state under the agreement between the Swedish government and the County Councils, the ALF-agreement (#ALFGBG-715986 and #ALFGBG-965240), the European Union Joint Program for Neurodegenerative Disorders (JPND2019-466-236), the Alzheimer’s Association 2021 Zenith Award (ZEN-21-848495), the Alzheimer’s Association 2022-2025 Grant (SG-23-1038904 QC), La Fondation Recherche Alzheimer (FRA), Paris, France, the Kirsten and Freddy Johansen Foundation, Copenhagen, Denmark, and Familjen Rönströms Stiftelse, Stockholm, Sweden.

Finally, the authors would like to thank all the current and past Neuropsychiatry Centre clinicians, and most importantly, patients and their families for their participation.

The corresponding author had full access to all the data in the study and had final responsibility for the decision to submit for publication.

## Statistical analysis conducted by

Dr Dhamidhu Eratne

