## Supplementary Material for "Diagnostic utility of plasma ptau217, ptau181, GFAP for Alzheimer disease in a heterogeneous younger onset dementia clinical cohort"

### Quantile regression results

A+T+ vs OtherAT

ptau217

Parameter	| Coefficient |	95% CI |	p

--------------------------------------------------------

(Intercept)	|	-0.51 | [-0.72, 0.10] | 0.108

csf status [A+T+] |	1.64 | [ 1.27, 2.14] | < .001

age	|	0.11 | [-0.01, 0.23] | 0.072

sex [Male]	|	-0.02 | [-0.72, 0.25] | 0.900

Model: scale(ptau217) ~ csf_status + scale(age) + sex (73 Observations)

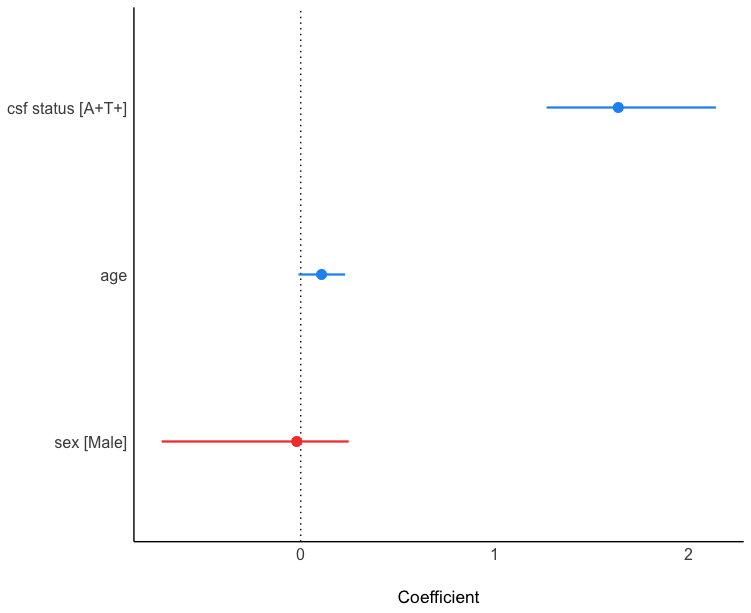

Ptau181

Parameter	| Coefficient |	95% CI |	p

--------------------------------------------------------

(Intercept)	|	-0.11 | [-0.65, 0.33] | 0.714

csf status [A+T+] |	1.07 | [ 0.18, 1.82] | 0.014

age	|	0.27 | [ 0.08, 0.47] | < .001

sex [Male]	|	-0.36 | [-0.91, 0.33] | 0.252

Model: scale(ptau181) ~ csf_status + scale(age) + sex (63 Observations)

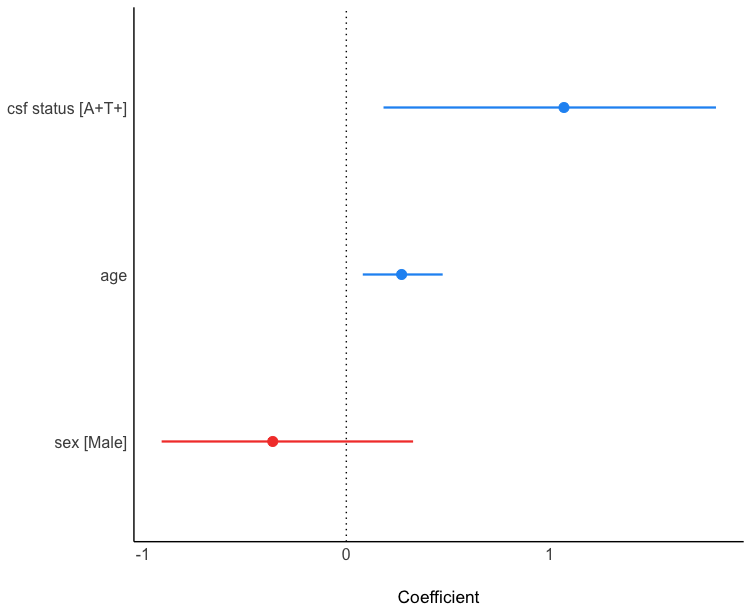

GFAP

Parameter	| Coefficient |	95% CI |	p

---------------------------------------------------------

(Intercept)	|	-0.43 | [-0.64, -0.16] | 0.004

csf status [A+T+] |	0.85 | [ 0.39, 1.35] | 0.006

age	|	0.19 | [ 0.07, 0.33] | < .001

sex [Male]	|	0.05 | [-0.30, 0.39] | 0.802

Model: scale(gfap) ~ csf_status + scale(age) + sex (79 Observations)

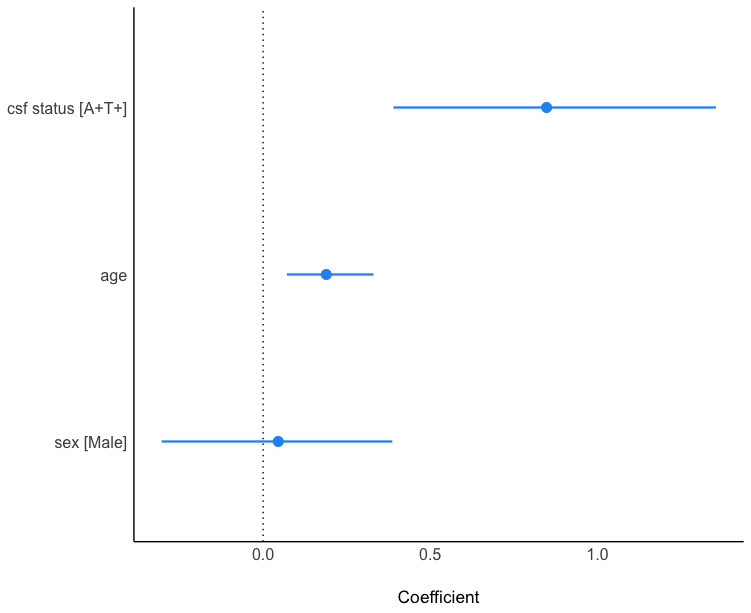

AB42:AB40

Parameter	| Coefficient |	95% CI |	p

-------------------------------------------------------

(Intercept)	|	0.52 | [-0.05, 0.91] | 0.074

csf status [A+T+] |	-0.57 | [-1.14, 0.10] | 0.098

age	|	-0.20 | [-0.39, 0.12] | 0.134

sex [Male]	|	-0.45 | [-0.98, 0.19] | 0.164

Model: scale(ab4240) ~ csf_status + scale(age) + sex (64 Observations)

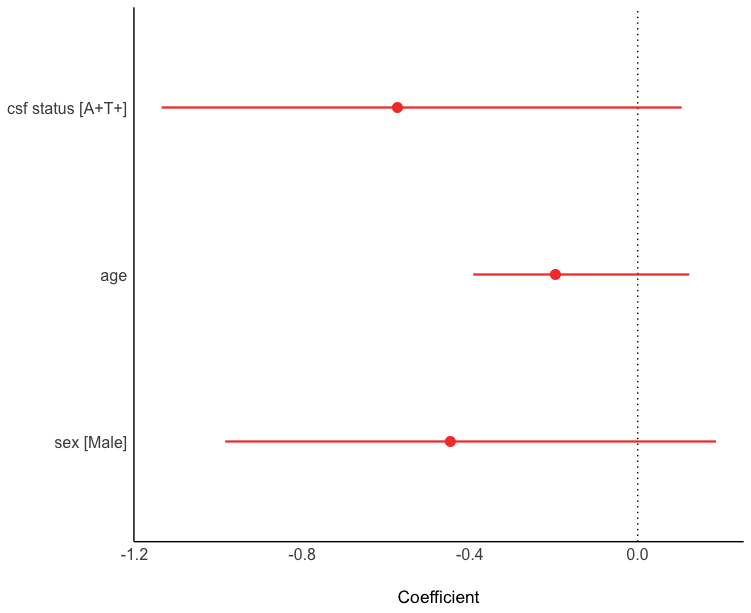

NfL

Parameter	| Coefficient |	95% CI |	p

---------------------------------------------------------

(Intercept)	|	-0.19 | [-0.38, -0.11] | 0.008

csf status [A+T+] |	0.02 | [-0.04, 0.32] | 0.302

age	|	0.02 | [ 0.01, 0.24] | < .001

sex [Male]	|	-6.05e-03 | [-0.23, 0.10] | 0.758

Model: scale(nfl) ~ csf_status + scale(age) + sex (79 Observations)

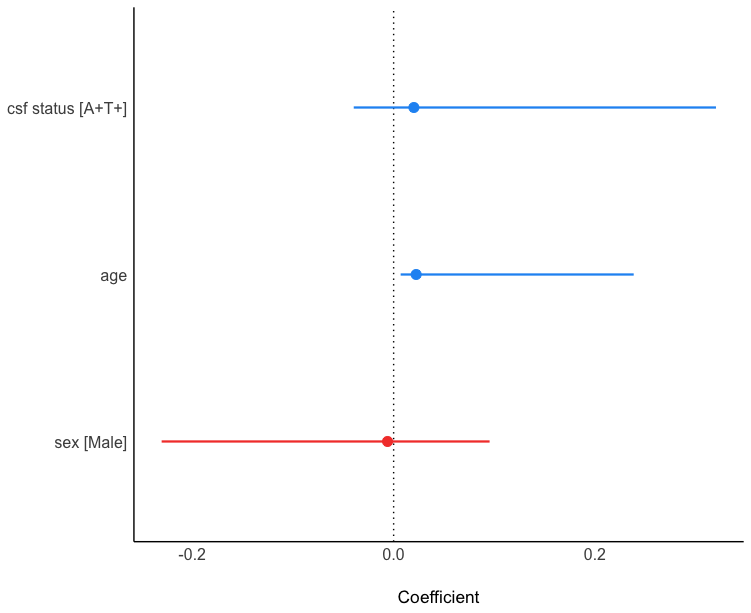

### Further details of ROC curve analyses

|  |  | Area under the curve | 95% CI | Optimal cut-off | Specificity | Sensitivity |
| --- | --- | --- | --- | --- | --- | --- |
| A+T+ vs OtherAT | ptau217 | 0.94 | 0.89, 0.99 | 2.35 | 88 | 100 |
|  | ptau181 | 0.79 | 0.65, 0.93 | 3.60 | 94 | 57 |
|  | GFAP | 0.72 | 0.58, 0.86 | 167 | 75 | 81 |
|  | AB42:AB40 | 0.65 | 0.52, 0.79 | 0.064 | 44 | 100 |
|  | NfL^NS^ | 0.56 | 0.44, 0.68 |  |  |  |
| A+T+ vs A-T- | ptau217 | 0.997 | 0.99, 1 | 2.06 | 96 | 100 |
|  | ptau181 | 0.87 | 0.74, 1 | 2.24 | 89 | 79 |
|  | GFAP | 0.81 | 0.66, 0.96 | 167 | 88 | 81 |
|  | AB42:AB40 | 0.46 | 0.58, 0.93 | 0.064 | 63 | 100 |
|  | NfL^NS^ | 0.51 | 0.33, 0.69 |  |  |  |
| A+T+ vs A+T- | ptau217 | 0.90 | 0.81, 1 | 2.35 | 83 | 100 |
|  | ptau181 | 0.77 | 0.61, 0.93 | 3.60 | 92 | 57 |
|  | GFAP^NS^ | 0.63 | 0.46, 0.80 |  |  |  |
|  | AB42:AB40^NS^ | 0.64 | 0.46, 0.81 |  |  |  |
|  | NfL^NS^ | 0.60 | 0.44, 0.76 |  |  |  |
| A+T+ vs A-T+ | ptau217 | 0.88 | 0.64, 1 | 2.06 | 80 | 100 |
|  | ptau181 | 0.56 | 0.21, 0.91 |  |  |  |
|  | GFAP | 0.78 | 0.50, 1 | 163.5 | 83 | 81 |
|  | AB42:AB40^NS^ | 0.67 | 0.36, 0.98 |  |  |  |
|  | NfL^NS^ | 0.57 | 0.19, 0.95 |  |  |  |

**Supplementary Table. Further details of ROC curve analyses**

NS: ROC curve analysis was not significant

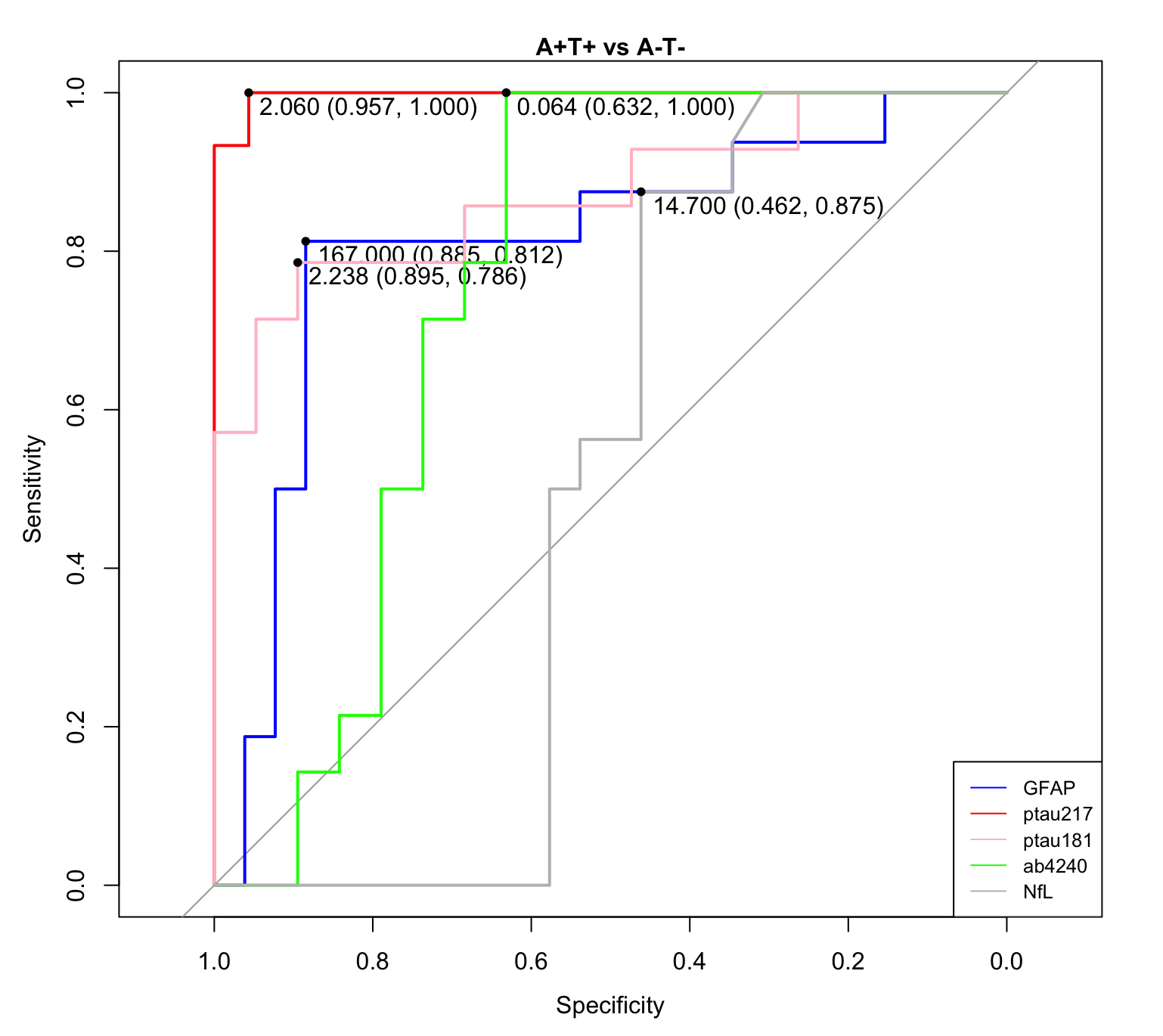

A+T+ vs A-T-

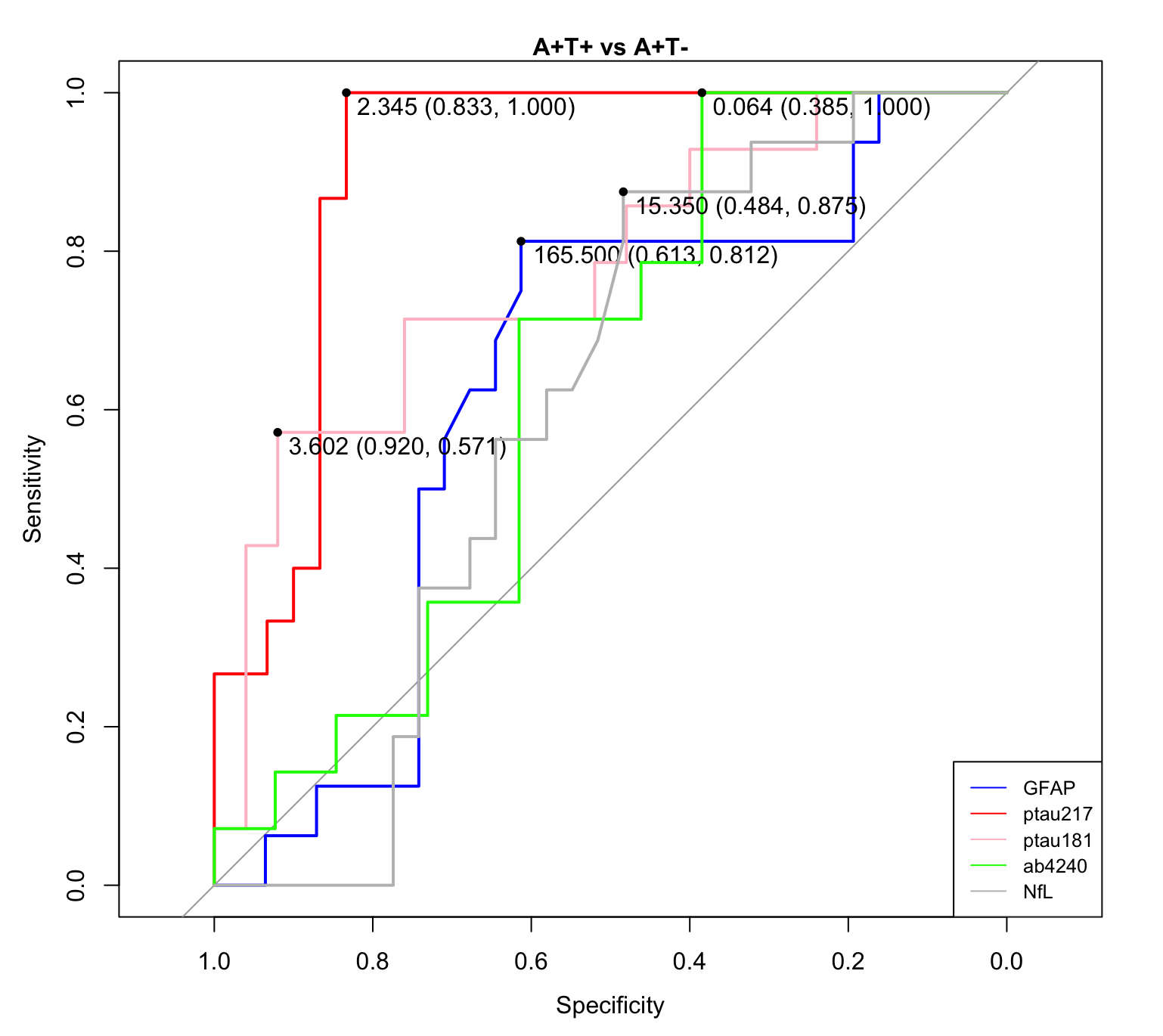

A+T+ vs A+T-

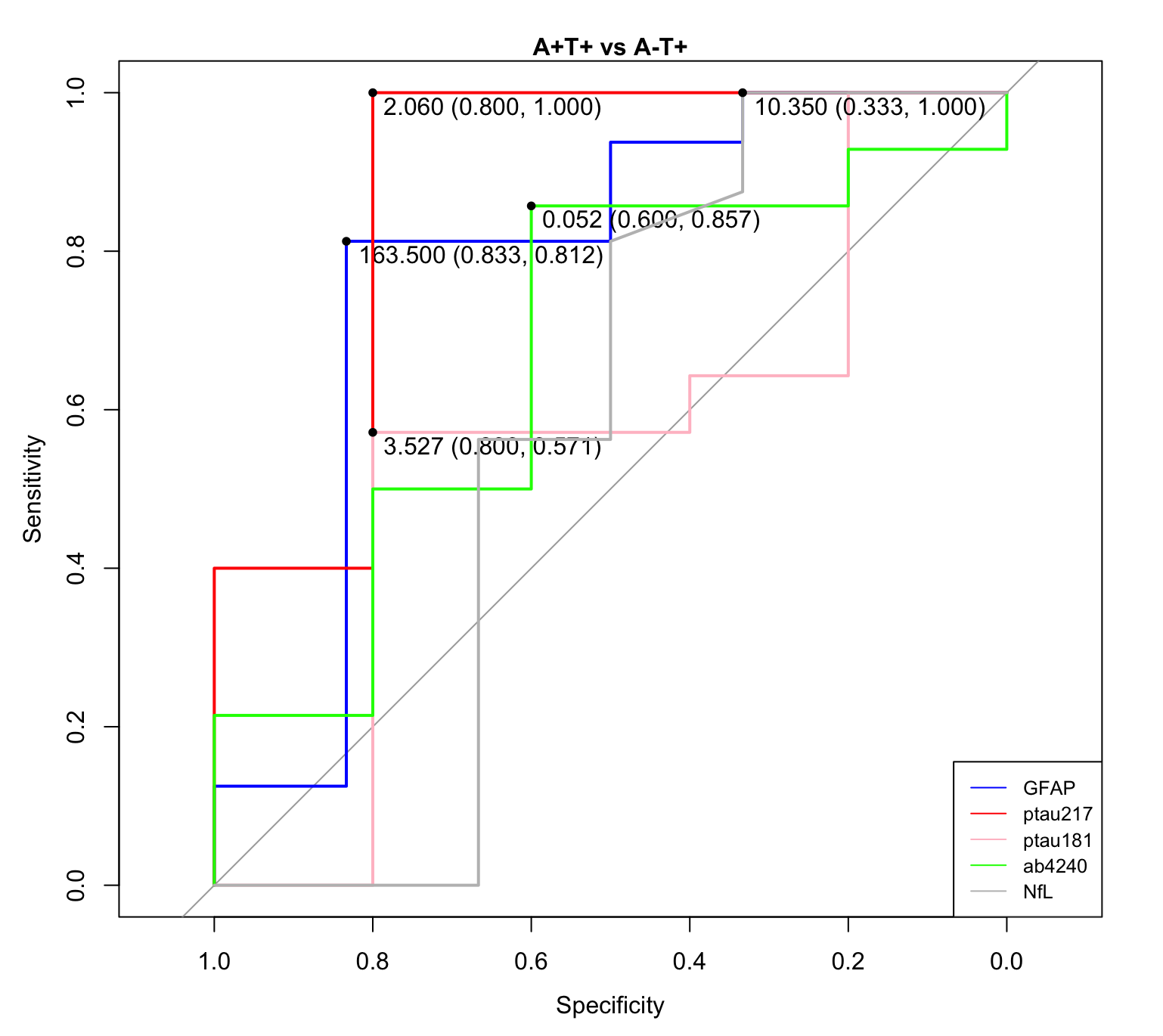

A+T+ vs A-T+
